# Structured Understanding of Assessment and Plans in Clinical Documentation

**DOI:** 10.1101/2022.04.13.22273438

**Authors:** Doron Stupp, Ronnie Barequet, I-Ching Lee, Eyal Oren, Amir Feder, Ayelet Benjamini, Avinatan Hassidim, Yossi Matias, Eran Ofek, Alvin Rajkomar

**Affiliations:** Google

## Abstract

Physicians record their detailed thought-processes about diagnoses and treatments as unstructured text in a section of a clinical note called the assessment and plan. This information is more clinically rich than structured billing codes assigned for an encounter but harder to reliably extract given the complexity of clinical language and documentation habits. We describe and release a dataset containing annotations of 579 admission and progress notes from the publicly available and de-identified MIMIC-III ICU dataset with over 30,000 labels identifying active problems, their assessment, and the category of associated action items (e.g. medication, lab test). We also propose deep-learning based models that approach human performance, with a F1 score of 0.88. We found that by employing weak supervision and domain specific data-augmentation, we could improve generalization across departments and reduce the number of human labeled notes without sacrificing performance.

## Introduction

After seeing a patient, physicians write a formal clinical note that records the patient ‘s history, findings, and conclusions. A key section, called the assessment and plan, is commonly written as a loosely structured problem-oriented list of conditions (e.g. “rheumatoid arthritis “), their assessments (e.g. “a new flare “), and plans (e.g. “CT of the neck and initiate treatment “). The text itself is free-form and written in physician-speciality-, and organizationally-idiosyncratic ways, making it hard to algorithmically parse into structured data. This structured data is valuable, for example, to drive a longitudinal visualization of when various diagnoses and treatments occurred. Relying on inherently structured data elements, such as coded ICD-9/10 diagnoses, can be unreliable^1^ and the free text is a more informative reflection of the physician ‘s assessment.

Prior research dealing with the conversion of the unstructured clinical note data into more structured forms has focused mostly on the entity matching and linking level (e.g. identifying diseases, drugs and their relations). Previous works have tackled various aspects of this problem, identifying entities^2–6^ and their relations^7,8^, identifying salient events^9^ and building personalized clinical knowledge graphs^10^. However, structuring full notes or sections remains mostly unexplored. The most closely related work is Mullenbach et al.^11^ which focused on identifying and classifying discharge instructions in discharge summaries, an important task for follow-up outpatient care after a hospitalization. In this work, we focus on inpatient assessment and plan sections and their richer problem-oriented structure, which includes not only action items (analogous to discharge instructions) but their associated condition and its assessment in admission and progress notes.

In this paper we describe a dataset and deep learning models for parsing an unstructured assessment and plan (A&P) section into segments indicating conditions, assessments, and plan action items. We present a dataset of 579 notes from the publicly available, de-identified, MIMIC-III ICU dataset^12^ with A&P sections annotated by clinicians for active problems and their associated assessment description and plan action items (more than 30,000 annotations). We then present a series of deep learning and hand-engineered models and analyze their performance. We show that models trained on this task approach human-comparable performance. In addition, training with weak supervision on clinically inspired heuristics followed by human labels as well as using data-augmentations provides the best performance. We then show these techniques can reduce the requirements for human labeled data and help the model generalize to notes written by other hospital departments.

## Results

At a high level, we had medical professionals annotate assessment and plan sections from a public dataset and trained a series of models to perform the same annotations using a variety of modeling techniques including weak supervision and data augmentation. We then examined how many manually annotated notes were necessary during training for good performance and generalization across notes from different hospital departments.

## Labeling and Data Collection

We classify spans of text in A&P sections related to the problem oriented structure into three categories: the active problem ‘s title (e.g. “sepsis “), it ‘s description assessment (e.g. “dd of UTI, pneumonia “), and plan action items (e.g. “trend WBC “). Text unrelated to these categories was left unlabelled. Action items were further broken down into eight different classes: medications, observations/labs, imaging, consults, nutrition, therapeutic procedures, other diagnostic procedures or other. Supplementary Figure 2 shows the frequency of each category. An example of an annotated note is shown in Supplementary Figure 1.

We randomly selected 579 A&P sections from physician-written notes from the MIMIC-III ICU dataset, which contains more than 50K patient stays from the Beth-Israel Deaconess medical center ICU wards^12^. Selected notes include both admission and progress notes from the ICU wards. Notes were sampled such that each patient was represented by at most one note. Approximately 90% of these notes were used for training and 10% for the test set. Unless indicated otherwise, all results are based on the test set.

Labels were generated with two procedures: human raters and clinical heuristics. Each note in the training set was labeled by one rater, the test set notes were labeled by six labelers, with a single physician acting as the ground truth (see methods).

Inter-rater agreement of the human raters measured as the Jaccard similarity was 0.77 (CI 0.75-0.79) for span type and 0.62 (CI 0.6-0.64) for action item type (both span level micro average, see Figure 1). While interrater agreement is high on average, there are differences in performance compared to the ground truth with span level micro average F1 scores ranging from 0.62 to 0.93. In contrast to the training set, the validation set was labeled by the raters with the highest performance (compared to the ground truth) leading to a potentially higher quality. For action item type classification, the confusion matrix (Supplementary Figure 4) shows most disagreement was found in the less frequent labels (“other diagnostic procedures” and “other “). Notably, disagreement was semantically congruent - e.g. labeling medications as therapeutic procedures. Qualitatively, these cases mostly involved edge-cases such as oxygen, blood products, fluids etc. in which the labeling instructions dictated the use of “medication” but raters opted for therapeutic procedures in many cases.

**Figure 1.**
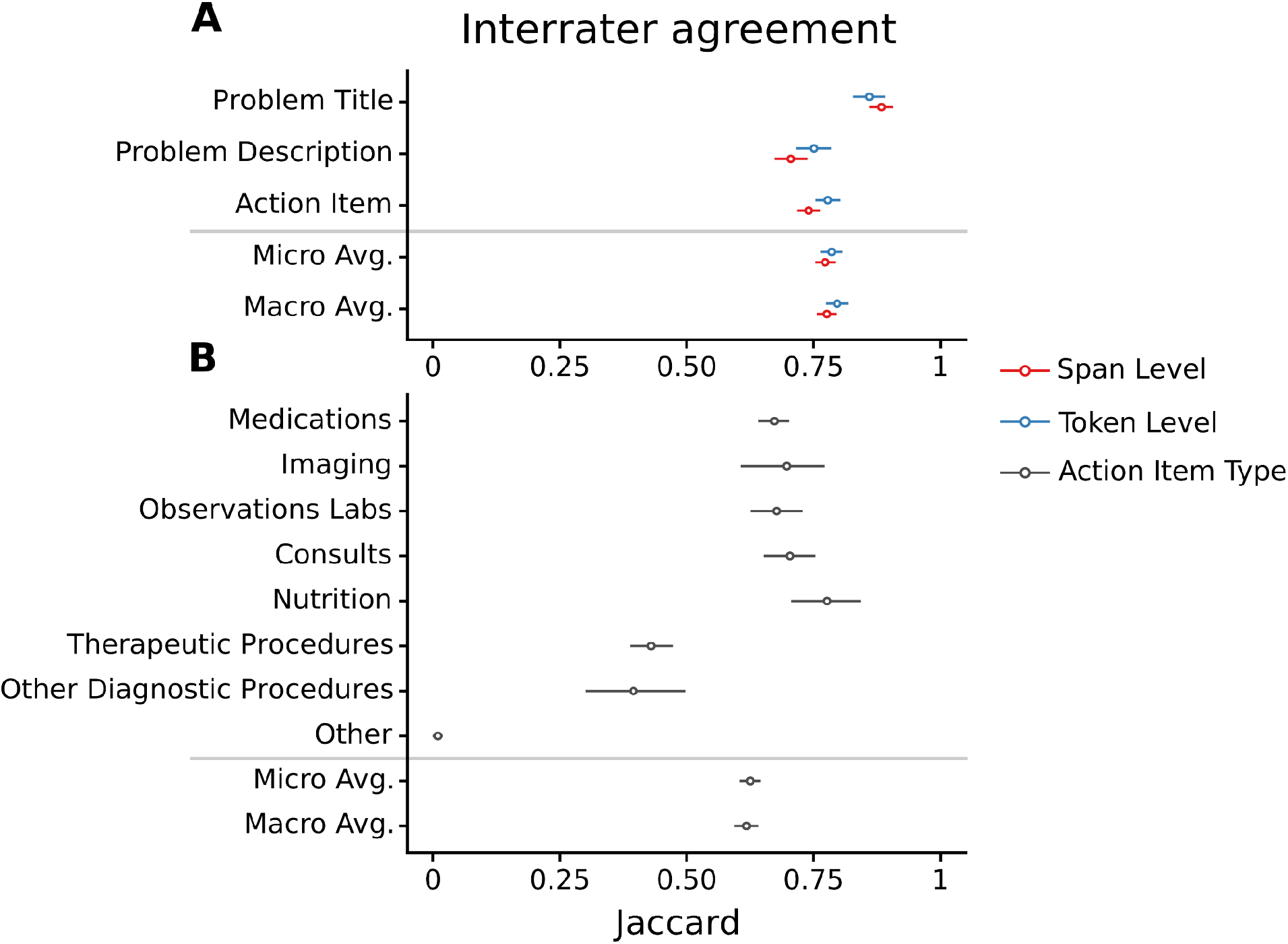
interrater agreement. Interater agreement measured as the average pairwise Jaccard index across raters. Agreement is measured across span types (A) and action item types (B). Span type Jaccard is presented at the span (red) and token (blue) level. Confidence intervals shown are the 95% percentile bootstrap intervals clustered across notes. Macro- and Micro- averages denote the simple and proportion weighted averages respectively.

The clinical heuristic was implemented using regular expressions to capture a bulleted list of active problems, followed by a bulleted list action items (see methods, Supplementary Figure 1 for examples). Importantly, the heuristic captures only the span type (problem title, description or action item). Action item type was not included in the clinical heuristic approach due to the lack of a simple heuristic to do so.

### The Model

We modeled the task of identifying the spans of active problem titles, descriptions and action items in the text as a sequence tagging task. The model consisted of a 2-layer bidirectional Long-short term memory network (LSTM)^13^ with a conditional random field (CRF) prediction head on top^14^. For each token, the model predicts its span type and action item type if applicable (see methods).

Since human-labeled data is expensive to obtain in the clinical domain, we sought to assess whether injecting domain knowledge into the model could increase data efficiency and potentially provide a boost in performance. Specifically, we focused on weak supervision and data augmentations. Both of these are known techniques in machine learning for improving model performance and lowering data-requirements and were explored for NLP and other tasks, including in the medical domain^15,16^.

To train the weakly supervised model, we used 25,000 notes that were labeled with the clinical heuristic described above. These notes were selected similarly to the human labeled notes, and belonged to a different set of patients, without overlap with notes already labeled by human raters. To implement data augmentations, we algorithmically restructured labeled assessment and plan sections (either human or heuristically labeled) to contain same-line action items, action items interleaved with descriptions and mixed bulleting (see examples in Supplementary Figure 5).

The models were trained in two phases, a “pre-training” phase with or without weak supervision, and with or without augmentations, and a second phase on non-augmented human labeled data only (see methods). To be specific, at the first phase, 4 models were trained for the 4 possible combinations of using weak supervision and/or data-augmentation. The models trained with weak supervision were pre-trained on the 25,000 random notes labeled with the clinical heuristic. For a fair comparison, models trained solely on the human labeled data were trained for the same overall number of steps, with a similar early stopping policy (see methods). As a baseline, we compared the performance of our models to that of the clinical heuristic. This provides a basic rule-based baseline for this new task.

All models reached significantly better performance than the baseline model, and approached the median rater in performance for span type (Table 1). Model loss patterns for span types are shown in Supplementary Figure 6. These loss patterns are qualitatively similar to the ones observed in the inter-rater comparison.

**Table 1.** Model performance. Model performance as F1 score across different training regimens. Aug - data augmentations. Macro- and micro-denote the simple and proportion weighted averages respectively. CI (95%) denotes the 95% percentile bootstrap confidence interval clustered across the notes. Numbers in brackets indicate the total number of spans in the test set. See supplementary table 2 for span and action item type detail.

|  | Action Item Type<br>(1063) |  |  |  |  |  | Span Level<br>(2016) |  |  |  |  |  | Token Level<br>(2016) |  |  |  |  |  |
| --- | --- | --- | --- | --- | --- | --- | --- | --- | --- | --- | --- | --- | --- | --- | --- | --- | --- | --- |
|  | Micro-F1 |  |  | Macro-F1 |  |  | Micro-F1 |  |  | Macro-F1 |  |  | Micro-F1 |  |  | Macro-F1 |  |  |
|  | Mean | CI (95%) |  | Mean | CI (95%) |  | Mean | CI (95%) |  | Mean | CI (95%) |  | Mean | CI (95%) |  | Mean | CI (95%) |  |
| Ratings | 0.744 | 0.705 | 0.778 | 0.807 | 0.778 | 0.834 | 0.847 | 0.822 | 0.870 | 0.841 | 0.817 | 0.864 | 0.853 | 0.819 | 0.883 | 0.860 | 0.832 | 0.885 |
| Ratings + aug | 0.757 | 0.726 | 0.787 | 0.806 | 0.782 | 0.830 | 0.866 | 0.842 | 0.888 | 0.862 | 0.836 | 0.885 | 0.862 | 0.832 | 0.888 | 0.872 | 0.845 | 0.895 |
| Pretrained | 0.734 | 0.698 | 0.769 | 0.786 | 0.755 | 0.815 | 0.849 | 0.827 | 0.870 | 0.843 | 0.823 | 0.863 | 0.854 | 0.825 | 0.880 | 0.863 | 0.840 | 0.885 |
| Pretrained + aug | 0.771 | 0.744 | 0.797 | 0.814 | 0.786 | 0.840 | 0.883 | 0.865 | 0.898 | 0.878 | 0.860 | 0.894 | 0.874 | 0.847 | 0.899 | 0.881 | 0.855 | 0.899 |
| Median Rater | 0.842 | 0.820 | 0.861 | 0.885 | 0.864 | 0.902 | 0.911 | 0.898 | 0.924 | 0.906 | 0.891 | 0.919 | 0.910 | 0.892 | 0.927 | 0.916 | 0.900 | 0.932 |
| Heuristic | - | - | - | - | - | - | 0.668 | 0.615 | 0.716 | 0.756 | 0.717 | 0.789 | 0.634 | 0.573 | 0.695 | 0.747 | 0.710 | 0.782 |

### Effect of training set size on model performance

We then studied model performance improvements when additional labeling is added as shown in Figure 2. Training with weak supervision and augmentations improved the performance on the span level across training dataset sizes, with performance converging when training on hundreds of notes. Importantly, a model trained on 25 notes with weak-supervision and augmentations (second data-point from the left in each panel) achieves comparable performance on the span level to a model trained with the full training data of 481 notes without weak supervision and no augmentations. However this effect is less pronounced at the token level. Notably, action item type labels are unavailable for the weak supervision as they ‘re not labeled by the heuristic, leading to a similar performance in both cases.

**Figure 2.**
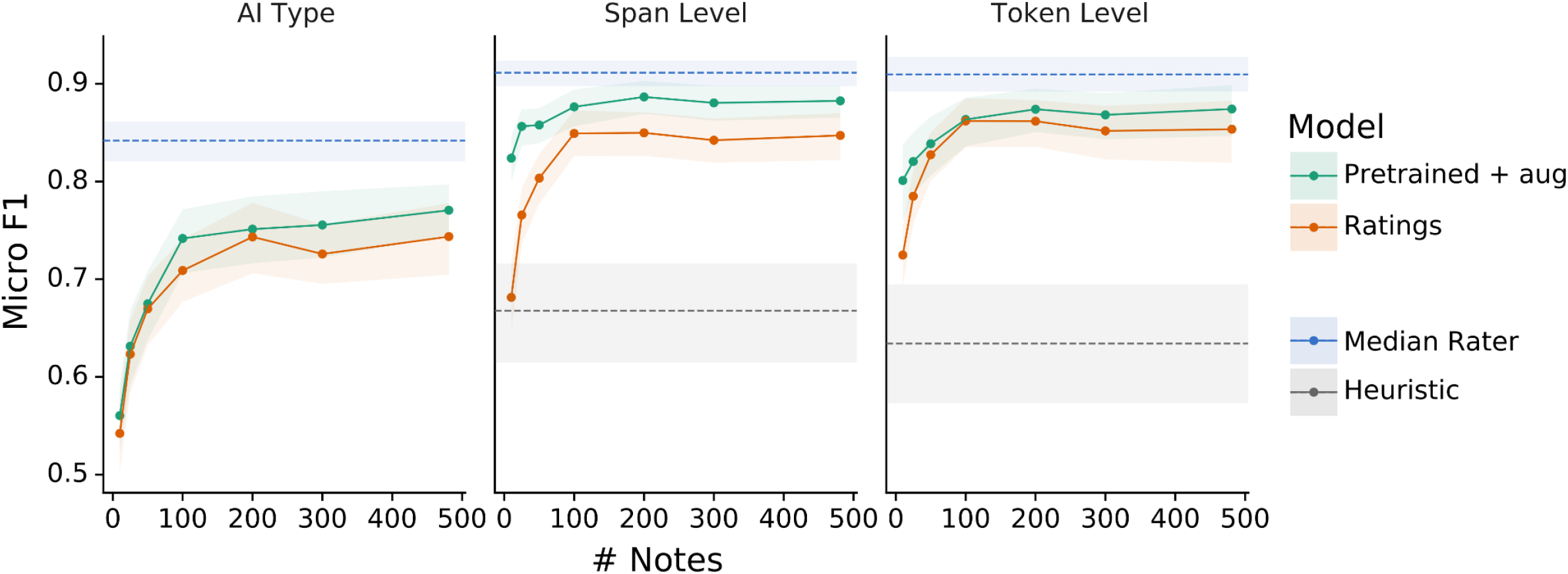
Performance with limited data. Model performance as micro-average F1 score across action item types (AI type, left), or span types at the span (middle) or token (right) level. Each dot represents the micro F1 score for the given model (by color) at the specific number of notes. Rightmost data points in all panels represent the full training data size (481 note). 95% percentile confidence intervals from clustered bootstrapping on the notes are represented as lighter shaded strips of the same color as their respective lines. Heuristic denotes the performance of the clinical heuristic (not available for action item type type, see methods).

### Generalization of performance across departments

Clinical models are often developed in a single center but deployed at other hospitals whose data distribution is unknown, making model generalization difficult to ascertain. To estimate this, we trained a series of models trained on a specific set of departments and tested them on different departments. We categorized notes by their respective service into two categories - medical or surgical (or other, see methods). The different services had a different distribution of spans (Supplementary Figure 3) and different typical note structures (see for example the notes in Supplementary Figure 1). Surgical notes tended to be formatted in a system oriented template, with roughly 30% of surgical notes having the exact same set of active problems. Descriptions and action items were commonly interleaved inline. Notes from medical services tended to be condition oriented with more variety in problem titles. Descriptions come mostly right after the problem title with a bulleted list of action items following (similar to the structure in the rule-based heuristic). We thus trained models on a single service and tested them on both services (Figure 3). Models trained with either augmentations or weak supervision tended to perform better both in-service and across-services. However, models trained on surgical notes and evaluated on medical notes had a significantly better performance on the span level when also trained with weak supervision. This may be attributed to the relatively high performance of the heuristic on notes from medical services.

**Figure 3.**
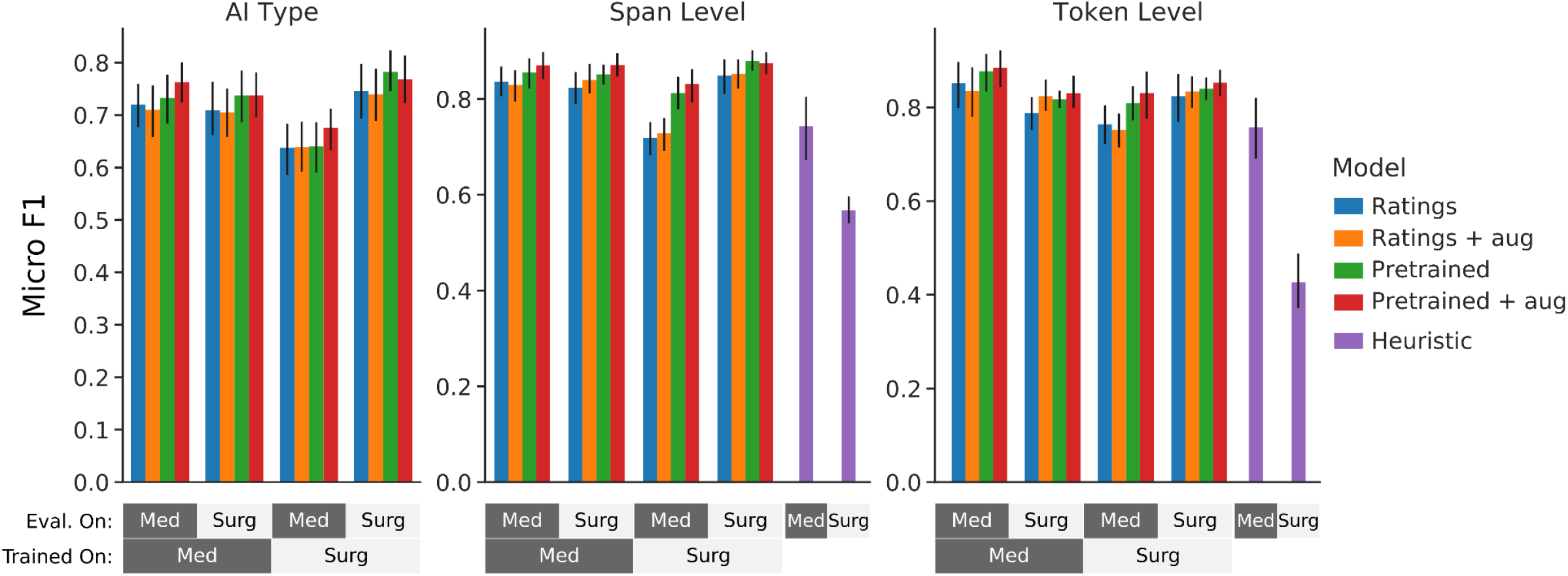
Generalization between departments. Model performance as micro average F1 scores trained on notes from one department (strip at the bottom) and evaluated on another (strip above). Model performance is compared when trained with or without weak supervision (pretraining) and with or without augmentations (abbreviated as aug). The clinical heuristic is presented for comparison (purple). Performance is measured for span type at the token level (right) and span level (middle) and for action item type (left, AI type). Error bars denote the 95% percentile confidence interval from clustered bootstrapping on the notes. Eval. on - evaluated on, Med - medical services, Surg - surgical services, AI type - action item type.

### Predictions Exploration

Structuring assessment and plan sections can uncover various aspects of patient care which are specified mostly or solely in the text. To address the potential utility of this structure, we generated predictions for all 141K physician notes in MIMIC-III, for a total of 4.7M predicted spans. We then explored the associated action items and descriptions for a series of common problems across (Table 2). Qualitatively, detected spans are indeed clinically relevant. For example, “cardiovascular” is associated with relevant medications (aspirin, beta blockers) and “respiratory failure” with relevant studies (chest x-ray, blood gases, cultures). Many of these spans may be difficult to associate with the problems with a more traditional relation-extraction approach as they do not correspond to clear entities (e.g. “avoid nephrotoxins “) or are ambiguous (“is” - incentive spirometer, “p.t.” - physiotherapy and patient).

**Table 2.**
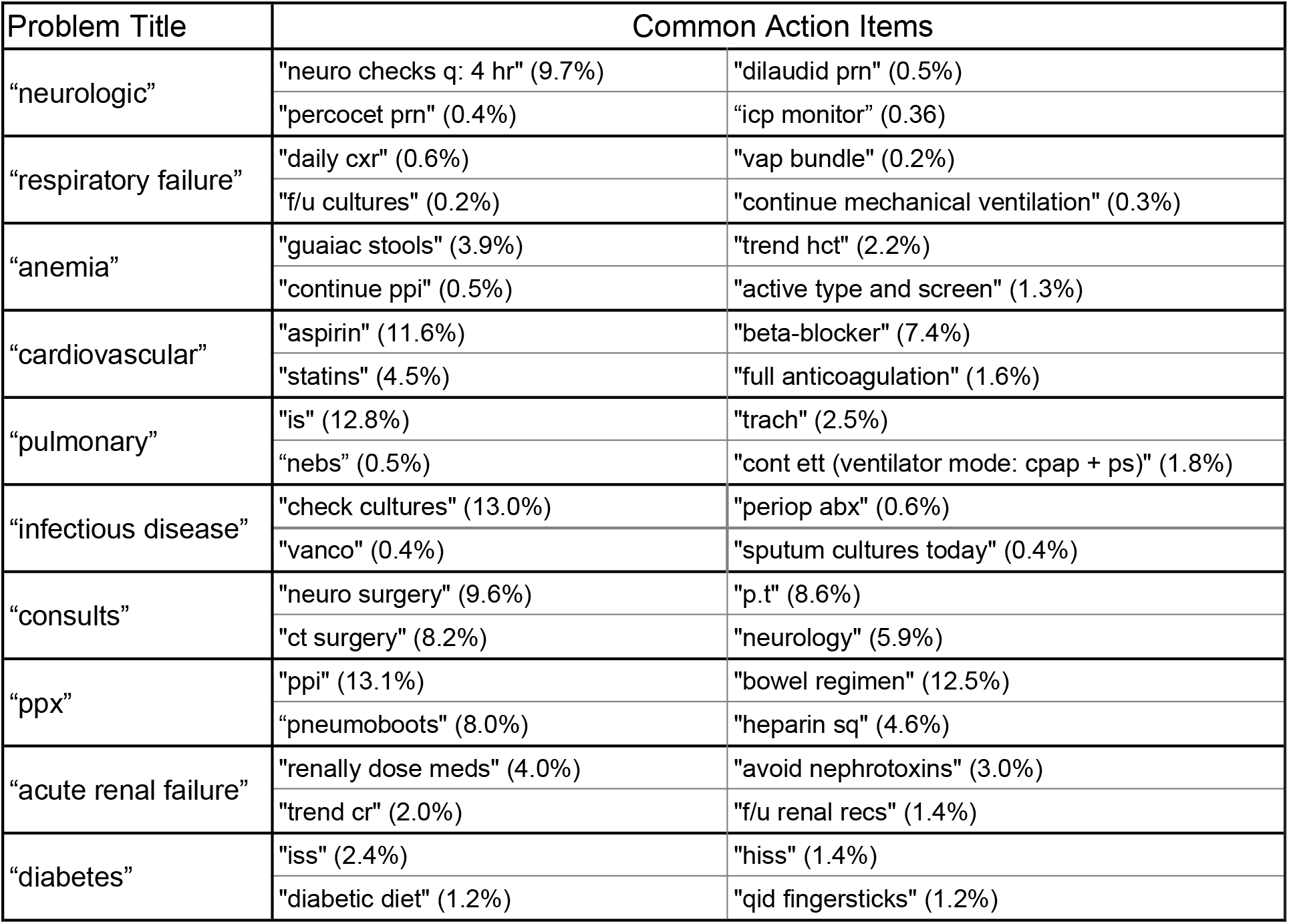
Common action items for common conditions. For each problem type (left column), common action items are presented (right columns). Percentage in brackets denote the percent of spans associated with the specified problem title mentioning this exact action item. Spans are presented “as-is” with no disambiguation or abbreviation expansion.

## Discussion

We demonstrate a method that can scalably and accurately parse unstructured clinician assessment and plan sections into structured data-elements without having to label a large number of notes by leveraging documentation domain-knowledge in the form of clinical heuristics and data-augmentation techniques. We also release a dataset of professionally labeled assessments and plans for the clinical community to improve on performance of machine learning models on this task.

Assessment and plan sections contain the main points of clinical thinking regarding the patient ‘s state. This includes information which may be otherwise hard to decipher from structured data such as labs or medication orders (i.e. the intent to obtain data). This task is inherently hard because documentation patterns are variable across institutions, departments and individual physicians. Clinician raters themselves had many disagreements on how to label notes, requiring non-trivial labeling instructions (Supplementary Text 1). Interestingly, we found the loss patterns of the model to be qualitatively similar to those found by inter-rater disagreement.

To achieve this performance, we utilized two approaches for supplementing the model with domain expertise - weak supervision derived from heuristics and data augmentations. Both approaches were previously used successfully in a number of domains^15^ including medical texts^16^. These approaches present an inherent time trade-off with human labeling. Supplementing with domain knowledge can be costly to develop, as the heuristics themselves are learnt from clinicians, while labeling time of medical experts may prove costly on its own. We show that supplementing the model with domain expertise can be beneficial for both the low data regime and across departments. This may prove useful when adapting a model for new hospital systems, generalizing to many new locations, where initial implementation is a one time cost unlike the recurring labeling needed.

This work ties into the recent research results to structure various notes and note parts such as discharge instructions^11^ and imaging reports^10^ to facilitate computational understanding of medical notes. Here we primarily focus on structuring progress in inpatient stays, however, a similar structure may be available for progress notes in the outpatient setting.

Our work has several limitations. A key finding of this work is that parsing the assessment and plan section itself has subjective elements, as highlighted by inter-rater disagreements. This leads to a limitation of the released dataset in which training and validation data is only labeled with a single rater. As performance is measured only on the test set, it may be the case that the training data is more heavily influenced by lower quality ratings leading to subpar model performance. As for the model, although the models presented in this paper may be improved further by utilizing recent advances in deep learning for natural language processing (e.g. by using transformers such as BERT^17^ or T5^18^), the smaller models presented here are commonly used and can be more easily deployed in a real-world EHR. Finally, generalizing across departments may not emulate generalization across locations properly. Some of the variance across notes is expected to arise from sources other than department practices - patient population, condition severity, frequency of care, EHR system-specific patterns and other unforeseen factors may all contribute to different documentation styles.

Future work could take advantage of the parsed assessment and plan to drive functionality to help clinicians understand a patient ‘s trajectory and history faster. For example, a temporal sequence of problems and their associated action items could be used, for example, to build timelines of patients ‘ trajectories of various conditions and treatments tried. Similarly, together with relation extraction, the extraction of problems and associated action items can be used to accurately build knowledge graphs across a population of patients.

In conclusion, we present models and an associated clinician-annotated dataset for structuring assessment and plan sections in clinician notes. We show that excellent performance can be achieved with a limited number of human-labeled notes, and maintained across departments, by incorporation of domain expertise in the form of weak supervision with clinically inspired heuristics and curated domain-specific data augmentations.

## Methods

### Labeling and Data Collection

Notes were sampled from the MIMIC-III^12^ note events table and filtered according to the category column to keep only physician notes. Notes were further manually inspected by the authors, keeping only notes with at least one non-synthetically generated assessment and plan section, discarding 2 notes in total. In total 579 notes were sampled and divided into a golden set of 48 notes, a training set of 481 notes and a validation set of 50 notes. The notes were labeled by 6 residents and medical students according to the labeling instructions as described in the results (and presented fully in Supplementary Text 1). Specific guidance was put forth to distinguish between descriptions and action items, splitting and merging spans and distinguishing between classes of action items.

Training and validation set notes were labeled by a single rater each. The golden set was labeled by all 6 raters with one of the raters (the primary author, rater id 1) acting as the ground truth. Rater 1 labeled the test set once before reviewing the other raters, which served as intra-rater comparison, and once after, which served as the golden set. Raters followed the labeling instructions found in Supplementary Text 1. Raters were evaluated before rating by completing either a small sample of notes first (and receiving feedback) or using the standardized quiz found in the instructions. The labels underwent automatic normalization to capture entire word boundaries and remove flanking non-alphanumeric characters. Intra-rater agreement was measured with approximately 2 months between a repeated attempt. Inter and Intra-rater agreement were calculated as the mean pairwise Jaccard index between raters.

### Service Attribution

Each note was associated with the service treating the patient at the time the note was written. Specifically, the service table was used and matched with the notes table on admission id, taking the most recent service per admission for the time of the note. Services were broadly categorized as “Medical “, “Surgical “, and “Other “. “Medical” includes “MED” (internal medicine), “CMED” (cardiology), “NMED” (neurology) and “OMED” (orthopedic medicine). “Surgical” includes “SURG” (general surgery), “TRAUM” (trauma), “CSURG” (cardiac surgery), “NSURG” (neurosurgery), “TSURG” (thoracic surgery), “VSURG” (vascular surgery), “ORTHO” (orthopedics), “ENT” (ear, nose and throat), “GU” (urology) and “GYN” (gynecology).

### Injecting domain expertise

Domain expertise was captured in the models in two ways: (i) weak supervision on a clinical heuristic and (ii) data augmentations. The heuristic was implemented using regular expression to capture active problems and their associated description and action items. This heuristic captures bulleted or numbered lists of active problems containing a bulleted list of action items (full python implementation can be found in the code). The heuristic is oblivious to action item types. Spans annotated with this heuristic serve as both a baseline model and labels for training the model with weak supervision as described below.

Data augmentations were designed to capture inline action items, action items interleaved with descriptions and mixed bulleting. A&P Sections were randomly selected to undergo data augmentations. Based on labels (heuristic or human labeled), sections were decomposed to the annotated spans and reconstructed according to the augmentation policy. Sections could undergo several separate sequences of augmentations which were all used as training data. Example augmentations are available in Supplementary Figure 5.

### Data Generation

Human labeled notes and 25,000 heuristic labeled notes were processed to generate the model ‘s train, validation and test sets. The 25,000 heuristic labeled notes were sampled similarly to the human labeled notes. Importantly, these sets share no overlap on the note or patient level. Moreover, notes in both sets are sampled such that each patient represented in the sample has only one note overall. The training set includes both heuristic based and human labels, the validation set and test set contain the rated validation and ground truth labeled golden set exclusively (i.e. no heuristic labels). The notes underwent extraction of their assessment and plan sections, a white-space based tokenization keeping tabs and line-breaks, data augmentation (as described above) and conversion into TensorFlow examples. A&P sections made up of synthetically generated text (e.g. ICD codes) were removed. Data augmentation was randomly applied to assessment and plan sections to generate sections with unique characteristics such as inconsistent bulleting, non-bulleted action items and interleaved problem descriptions and action items (see Supplementary Figure 5 for examples). Data augmentations were applied to both human labeled and heuristic labeled notes. Augmentations were randomly sampled to yield 2 augmented views per note on average (poisson distributed). Augmented views were fed to the model in addition to the original note.

### Model

The model consists of a multilayer bidirectional LSTM with a CRF head on top. Token id embeddings were initialized from Word2Vec embeddings trained on Wikipedia^19^ (link: https://tfhub.dev/google/Wiki-words-250-with-normalization/2). The model is trained with a sequence tagging objective similar to named entity recognition models. It predicts for each token whether it belongs to a span (problem title, description or action item) and for action items, the action item type (medication, observations/labs, imaging, consults, nutrition, therapeutic procedures, other diagnostic procedures or other). Span labels are encoded as IOB2, action items labels are encoded as categoricals. The model is then trained with a CRF negative log likelihood loss for both heads (span type and action item type). The model was implemented using the TensorFlow^20^ 2 Keras API and the TensorFlow model garden^21^. For inference, span types are taken from the Viterbi decoding of the span type CRF head, action item types are predicted as the maximal likelihood type across the predicted span calculated from the logits (equivalent to Viterbi decoding with a diagonal transition matrix). For more information see the source code.

All models were trained with the same hyperparameters and the same training regimen. Briefly, the hyperparameters are an embedding size of 250, an LSTM hidden dimension of 256 (per direction), a learning rate of 1e-3 with linear decay. The regimen consists of two phases, in the first phase the models are pre-trained with either the weak supervision on heuristic derived labels or human labels, with or without augmentations for 2000 steps. For the second phase, the models are trained for 500 additional steps on labeled data without augmentations with a constant learning rate of 5e-4. In the second phase, early stopping is performed based on the macro average token level accuracy for both span and action item types.

### Evaluation

Models and ratings were evaluated on the span, token and action item type levels. At the span level, spans were considered correct if any overlap is found between a predicted and ground truth span. Every span can only match one other span, the largest overlapping span is considered the match. On the token level, every non-space token is considered correct if matching on the span type. For action item type, types are considered correct if spans match (via the span criterion) and the predicted action item type is the same as the ground truth. Then, precision, recall, Jaccard and F1 scores are calculated. 95% percentile confidence intervals for the metrics were calculated using the clustered bootstrap method, clustering on the note level.

## Data Availability

All data produced in the present work are either contained in the manuscript or available online on Zenodo at: https://doi.org/10.5281/zenodo.6413405

## Acknowledgements

The authors would like to thank Yun Liu, Aviel Atias, Doron Sharabani and Eyal Marcus for their helpful discussions during the project and their comments on the manuscript. The authors are employed by Google, LLC and own equity in Alphabet, Inc.

## Code Availability

Code is available on GitHub at: https://github.com/google-research/google-research/tree/master/assessment_plan_modeling under the Apache 2.0 license. The repository contains the code required to generate TensorFlow examples from notes and ratings, and train models against the ground truth.

## Data Availability

Annotations are available on Zenodo at: https://doi.org/10.5281/zenodo.6413405. Annotations are available as a CSV containing a single annotated span per row. Each row contains the original note row id (from the MIMIC-III note event table), character indices of the span, span and action item type and a rater unique id. Annotation stratification is denoted for training, validation and the test set. The test set is further divided into ground truth and other raters.

