## Supplementary Materials for "Structured Understanding of Assessment and Plans in Clinical Documentation"

Supplementary materials for “Structured Understanding of Assessment and Plans in Clinical Documentation” by Stupp et al.

Supplementary Table 1 - Data properties

| Span Type | # Labels | Frequency |
|---|---|---|
| Problem Title | 9475 | 30.3% |
| Problem Description | 6093 | 19.5% |
| Action Item | 15706 | 50.2% |
| Medications | 6915 | 44.0% |
| Observations/Labs | 3423 | 21.8% |
| Therapeutic Procedures | 2079 | 13.2% |
| Consults | 1297 | 8.3% |
| Nutrition | 885 | 5.6% |
| Imaging | 704 | 4.5% |
| Other Diagnostic Procedures | 305 | 1.9% |
| Other | 98 | 0.6% |
| **Total** | **31274** | |

Number and frequency of the various annotations. Action item types are presented below the action item span type, their frequency represent percent of action item spans.

#### Supplementary Table 2 - F1 Scores for Span and Action Item Types

| Span Type | Span Level | | | | | | | | | Token Level | | | | | | | | |
|---|---|---|---|---|---|---|---|---|---|---|---|---|---|---|---|---|---|---|
| | **Problem Title** (589) | | | **Problem Description** (364) | | | **Action Item** (1063) | | | **Problem Title** (589) | | | **Problem Description** (364) | | | **Action Item** (1063) | | |
| | Mean | CI (95%) | | Mean | CI (95%) | | Mean | CI (95%) | | Mean | CI (95%) | | Mean | CI (95%) | | Mean | CI (95%) | |
| Ratings | 0.941 | 0.920 | 0.961 | 0.759 | 0.721 | 0.797 | 0.823 | 0.791 | 0.852 | 0.930 | 0.893 | 0.962 | 0.809 | 0.758 | 0.854 | 0.841 | 0.802 | 0.874 |
| Ratings + aug | 0.957 | 0.924 | 0.982 | 0.784 | 0.740 | 0.825 | 0.844 | 0.816 | 0.870 | 0.943 | 0.904 | 0.976 | 0.822 | 0.774 | 0.862 | 0.850 | 0.814 | 0.880 |
| Pretrained | 0.936 | 0.914 | 0.955 | 0.767 | 0.730 | 0.805 | 0.827 | 0.798 | 0.853 | 0.932 | 0.901 | 0.957 | 0.823 | 0.782 | 0.859 | 0.834 | 0.798 | 0.867 |
| Pretrained + aug | 0.958 | 0.940 | 0.973 | 0.820 | 0.787 | 0.853 | 0.856 | 0.836 | 0.874 | 0.952 | 0.927 | 0.973 | 0.833 | 0.790 | 0.871 | 0.857 | 0.823 | 0.886 |
| Median Rater | 0.995 | 0.990 | 0.999 | 0.838 | 0.810 | 0.864 | 0.884 | 0.861 | 0.904 | 0.983 | 0.965 | 0.996 | 0.865 | 0.833 | 0.896 | 0.900 | 0.877 | 0.922 |
| Heuristic | 0.934 | 0.891 | 0.968 | 0.627 | 0.575 | 0.676 | 0.709 | 0.643 | 0.767 | 0.860 | 0.807 | 0.908 | 0.642 | 0.577 | 0.706 | 0.783 | 0.712 | 0.839 |

| Action Item Type | **Medications** (491) | | | **Imaging** (42) | | | **Observations Labs** (211) | | | **Consults** (86) | | | **Nutrition** (51) | | | **Therapeutic Procedures** (167) | | | **Other Diagnostic Procedures** (15) | | |
|---|---|---|---|---|---|---|---|---|---|---|---|---|---|---|---|---|---|---|---|---|---|
| | Mean | CI (95%) | | Mean | CI (95%) | | Mean | CI (95%) | | Mean | CI (95%) | | Mean | CI (95%) | | Mean | CI (95%) | | Mean | CI (95%) | |
| Ratings | 0.749 | 0.703 | 0.787 | 0.903 | 0.824 | 0.970 | 0.794 | 0.740 | 0.843 | 0.910 | 0.854 | 0.957 | 0.925 | 0.876 | 0.968 | 0.653 | 0.584 | 0.725 | 0.833 | 0.667 | 1.000 |
| Ratings + aug | 0.780 | 0.741 | 0.814 | 0.860 | 0.783 | 0.935 | 0.804 | 0.758 | 0.846 | 0.918 | 0.869 | 0.962 | 0.917 | 0.864 | 0.965 | 0.621 | 0.563 | 0.678 | 0.889 | 0.667 | 1.000 |
| Pretrained | 0.754 | 0.717 | 0.791 | 0.816 | 0.729 | 0.907 | 0.771 | 0.718 | 0.820 | 0.878 | 0.822 | 0.932 | 0.925 | 0.873 | 0.968 | 0.636 | 0.560 | 0.710 | 0.917 | 0.667 | 1.000 |
| Pretrained + aug | 0.806 | 0.775 | 0.836 | 0.895 | 0.819 | 0.967 | 0.805 | 0.756 | 0.856 | 0.888 | 0.836 | 0.939 | 0.908 | 0.849 | 0.962 | 0.650 | 0.580 | 0.717 | 0.867 | 0.667 | 1.000 |
| Median Rater | 0.877 | 0.847 | 0.908 | 0.988 | 0.970 | 1.000 | 0.906 | 0.877 | 0.935 | 0.921 | 0.883 | 0.956 | 0.947 | 0.906 | 0.982 | 0.672 | 0.600 | 0.742 | 0.941 | 0.861 | 1.000 |

Model performance as F1 score across different training regimens. Aug - data augmentations, AI type - action item type. CI (95%) denotes the 95% percentile bootstrap confidence interval clustered across the notes. Categories are denoted by type with total number of spans in the test set at brackets.

#### Supplementary Table 3 - All Performance Metrics

Attached as a separate file. Contains model performance as precision, recall, jaccard, F1 score and their sub components (true positives, total positives, total predicted) across different training regimens. Mu denotes the mean, CI (95%) denotes the 95% percentile bootstrap confidence interval clustered across the notes. Description in brackets denote the data the specific model was trained on (e.g. Pretrained+Augmentations (100 notes) is the model trained on 100 notes from the trainings data). The file contains 2 tabs, one for comparisons with the full test set, and one by service.

Supplementary Figure 1 - Example Notes

### A Ground Truth

Assessment and Plan:

Neurologic: d/c epidural Med.
Cardiovascular: stable, off neo
Pulmonary: Cont ETT, (Ventilator mode: CPAP + PS) Therap. Therap.
Gastrointestinal / Abdomen: Place NGT Therap.
Nutrition: npo Nut.
Renal: Foley, Adequate UO Therap.
Hematology:
Endocrine: RISS Med.
Infectious Disease: Therap. Therap.
Lines / Tubes / Drains: Foley, Chest tube - pleural
Wounds: Dry dressings Therap.
Imaging: CXR today Imag.
Fluids: LR Med.
Consults: thoracic Cons.

### B Heuristic

Assessment and Plan:

Neurologic: d/c epidural
Cardiovascular: stable, off neo
Pulmonary: Cont ETT, (Ventilator mode: CPAP + PS)
Gastrointestinal / Abdomen: Place NGT
Nutrition: npo
Renal: Foley, Adequate UO
Hematology:
Endocrine: RISS
Infectious Disease:
Lines / Tubes / Drains: Foley, Chest tube - pleural
Wounds: Dry dressings
Imaging: CXR today
Fluids: LR
Consults: thoracic

#### C Model

Assessment and Plan:

Neurologic: d/c epidural Med.
Cardiovascular: stable, off neo
Pulmonary: Cont ETT, (Ventilator mode: CPAP + PS) Therap.
Gastrointestinal / Abdomen: Place NGT Therap.
Nutrition: npo Nut.
Renal: Foley, Adequate UO Therap.
Hematology:
Endocrine: RISS Med.
Infectious Disease:
Lines / Tubes / Drains: Foley, Chest tube - pleural
Wounds: Dry dressings Therap.
Imaging: CXR today Imag.
Fluids: LR Med.
Consults: thoracic Cons.

### E Heuristic

Assessment and Plan
64 M w/h/o colon cancer s/p resection, ESRD on HD, and ischemic cardiomyopathy who was transferred to [**Hospital1 54**] for further evaluation of right hip fracture on [**6-26**], transferred to [**Hospital Unit Name 44**] with hypercarbic and hypoxic respiratory failure.

### Acute on Chronic hypercarbic and hypoxic respiratory failure:

- patient is much improved, likely due to narcotics and CT contrast dye causing pulmonary edema. 3 L fluid removed w/ HD yesterday with marked improvement in symptoms
- -still fluid overloaded on exam significantly, will need to slowly remove fluid over time with dialysis, patient is anuric.
- CPAP at night.

### Altered mental status: improved, likely due to narcotics / CO2 retention.

### Pain: continue Oxycontin at home dose 20 mg [**Hospital1 **] and prn oxycodone, 5mg po q4hr prn.

### R Hip Fx/RLE Weakness, - pathologic fractures of right iliac crest and right capital femoral head since 2/[**2177**]. Unclear if this is related to metastasis versus new primary. s/p biopsy at OSH, sent to B+W for further eval.

-oncology has discussed w/ ortho onc, ortho onc plan to biopsy when patient more stable

-SPEP reveals non specific bence [**Doctor Last Name **] protein, lamda type, free kappa / lamda sent, per onc may need bone marrow biopsy

### ESRD- on HD MWF-

- continue HD, cont nephrocaps, Ca acetate

### Afib- on tele, LFTs normal while on amiodarone, may not need amio if in chronic a fib but if paroxysmal then amio may reduce his a fib burden.

- cont amiodarone, metoprolol, diltiaze

### Ischemic cardiomyopathy: continue aspirin, metoprolol and statin.

-ruled out for MI.

### Gout - cont renally-dosed allopurinol

### Colon Cancer- defer to primary oncologist

### DM- cont humalog ISS

### PPx: cont PPI, sc heparin

### FEN: Renal Diet as tolerated, monitor lytes

### CODE: Full Code (confirmed with wife on transfer)

### F Model

Assessment and Plan
64 M w/h/o colon cancer s/p resection, ESRD on HD, and ischemic cardiomyopathy who was transferred to [**Hospital1 54**] for further evaluation of right hip fracture on [**6-26**], transferred to [**Hospital Unit Name 44**] with hypercarbic and hypoxic respiratory failure.
### Acute on Chronic hypercarbic and hypoxic respiratory failure:
- patient is much improved, likely due to narcotics and CT contrast dye causing pulmonary edema. 3 L fluid removed w/ HD yesterday with marked improvement in symptoms
- -still fluid overloaded on exam significantly, will need to slowly remove fluid over time with dialysis, patient is anuric.
- CPAP at night. Therap.
### Altered mental status: improved, likely due to narcotics / CO2 retention.
### Pain: continue Oxycontin at home dose 20 mg [**Hospital1 **] and prn oxycodone, 5mg po q4hr prn. Med.
### R Hip Fx/RLE Weakness, - pathologic fractures of right iliac crest and right capital femoral head since 2/[**2177**]. Unclear if this is related to metastasis versus new primary. s/p biopsy at OSH, sent to B+W for further eval.
-oncology has discussed w/ ortho onc, ortho onc plan to biopsy when Cons. patient more stable Cons.
-SPEP reveals non specific bence [**Doctor Last Name **] protein, lamda type, free kappa / lamda sent, per onc may need bone marrow biopsy Obs./Labs
### ESRD- on HD MWF-
- continue HD, cont nephrocaps, Ca acetate Med. Med.
### Afib- on tele, LFTs normal while on amiodarone, may not need amio if Med. in chronic a fib but if paroxysmal then amio may reduce his a fib burden.
- cont amiodarone, metoprolol, diltiaze Med.
### Ischemic cardiomyopathy: continue aspirin, metoprolol and statin. Med.
-ruled out for MI.
### Gout - cont renally-dosed allopurinol Med.
### Colon Cancer- defer to primary oncologist Cons.
### DM- cont humalog ISS Med.
### PPx: cont PPI, sc heparin Med. Med.
### FEN: Renal Diet as tolerated, monitor lytes Nut. Obs./Labs
### CODE: Full Code (confirmed with wife on transfer)

#### F Model

Assessment and Plan

64 M w/h/o colon cancer s/p resection, ESRD on HD, and ischemic cardiomyopathy who was transferred to [**Hospital1 54**] for further evaluation of right hip fracture on [**6-26**], transferred to [**Hospital Unit Name 44**] with hypercarbic and hypoxic respiratory failure.

### Acute on Chronic hypercarbic and hypoxic respiratory failure:

- patient is much improved, likely due to narcotics and CT contrast dye causing pulmonary edema. 3 L fluid removed w/ HD yesterday with marked improvement in symptoms

- -still fluid overloaded on exam significantly, will need to slowly remove fluid over time with dialysis, patient is anuric.

- CPAP at night. Therap.

### Altered mental status: improved, likely due to narcotics / CO2 retention.

### Pain: continue Oxycontin at home dose 20 mg [**Hospital1 **] and prn oxycodone, 5mg po q4hr prn. Med.

### R Hip Fx/RLE Weakness, - pathologic fractures of right iliac crest and right capital femoral head since 2/[**2177**]. Unclear if this is related to metastasis versus new primary. s/p biopsy at OSH, sent to B+W for further eval.

-oncology has discussed w/ ortho onc, ortho onc plan to biopsy when Cons. patient more stable Cons.

-SPEP reveals non specific bence [**Doctor Last Name **] protein, lamda type, free kappa / lamda sent, per onc may need bone marrow biopsy Obs./Labs

### ESRD- on HD MWF-

- continue HD, cont nephrocaps, Ca acetate Med. Med.

### Afib- on tele, LFTs normal while on amiodarone, may not need amio if Med. in chronic a fib but if paroxysmal then amio may reduce his a fib burden.

- cont amiodarone, metoprolol, diltiaze Med.

### Ischemic cardiomyopathy: continue aspirin, metoprolol and statin. Med.

-ruled out for MI.

### Gout - cont renally-dosed allopurinol Med.

### Colon Cancer- defer to primary oncologist Cons.

### DM- cont humalog ISS Med.

### PPx: cont PPI, sc heparin Med. Med.

### FEN: Renal Diet as tolerated, monitor lytes Nut. Obs./Labs

### CODE: Full Code (confirmed with wife on transfer)

### F Model

Assessment and Plan
64 M w/h/o colon cancer s/p resection, ESRD on HD, and ischemic cardiomyopathy who was transferred to [**Hospital1 54**] for further evaluation of right hip fracture on [**6-26**], transferred to [**Hospital Unit Name 44**] with hypercarbic and hypoxic respiratory failure.

### Acute on Chronic hypercarbic and hypoxic respiratory failure:
- patient is much improved, likely due to narcotics and CT contrast dye causing pulmonary edema. 3 L fluid removed w/ HD yesterday with marked improvement in symptoms
- -still fluid overloaded on exam significantly, will need to slowly remove fluid over time with dialysis, patient is anuric.
- CPAP at night. Therap.

### Altered mental status: improved, likely due to narcotics / CO2 retention.

### Pain: continue Oxycontin at home dose 20 mg [**Hospital1 **] and prn oxycodone, 5mg po q4hr prn. Med.

### R Hip Fx/RLE Weakness, - pathologic fractures of right iliac crest and right capital femoral head since 2/[**2177**]. Unclear if this is related to metastasis versus new primary. s/p biopsy at OSH, sent to B+W for further eval.
-oncology has discussed w/ ortho onc, ortho onc plan to biopsy when Cons. patient more stable Cons.
-SPEP reveals non specific bence [**Doctor Last Name **] protein, lamda type, free kappa / lamda sent, per onc may need bone marrow biopsy Obs./Labs

### ESRD- on HD MWF-
- continue HD, cont nephrocaps, Ca acetate Med. Med.

### Afib- on tele, LFTs normal while on amiodarone, may not need amio if Med. in chronic a fib but if paroxysmal then amio may reduce his a fib burden.
- cont amiodarone, metoprolol, diltiaze Med.

### Ischemic cardiomyopathy: continue aspirin, metoprolol and statin. Med.
-ruled out for MI.

### Gout - cont renally-dosed allopurinol Med.

### Colon Cancer- defer to primary oncologist Cons.

### DM- cont humalog ISS Med.

### PPx: cont PPI, sc heparin Med. Med.

### FEN: Renal Diet as tolerated, monitor lytes Nut. Obs./Labs

### CODE: Full Code (confirmed with wife on transfer)

Example notes from surgical (A-C) and medical (D-F) services. Spans from the ground truth (A, D), clinical heuristic (B, E) and model (C, F) are highlighted. Spans display the active problems (purple), descriptions (blue) and action items (green). Ovals denote the action item type. Action item types are shown for the same line action items or immediately below, in order from left to right. Action item type abbreviations used: Med. - medications, Imag. - imaging, Obs./Labs - observations / labs, Cons. - consults, Nut. - nutrition, Therap. - therapeutic procedures, Diag. - other diagnostic procedures.

### Supplementary Figure 2 - Span Type per Note Distribution

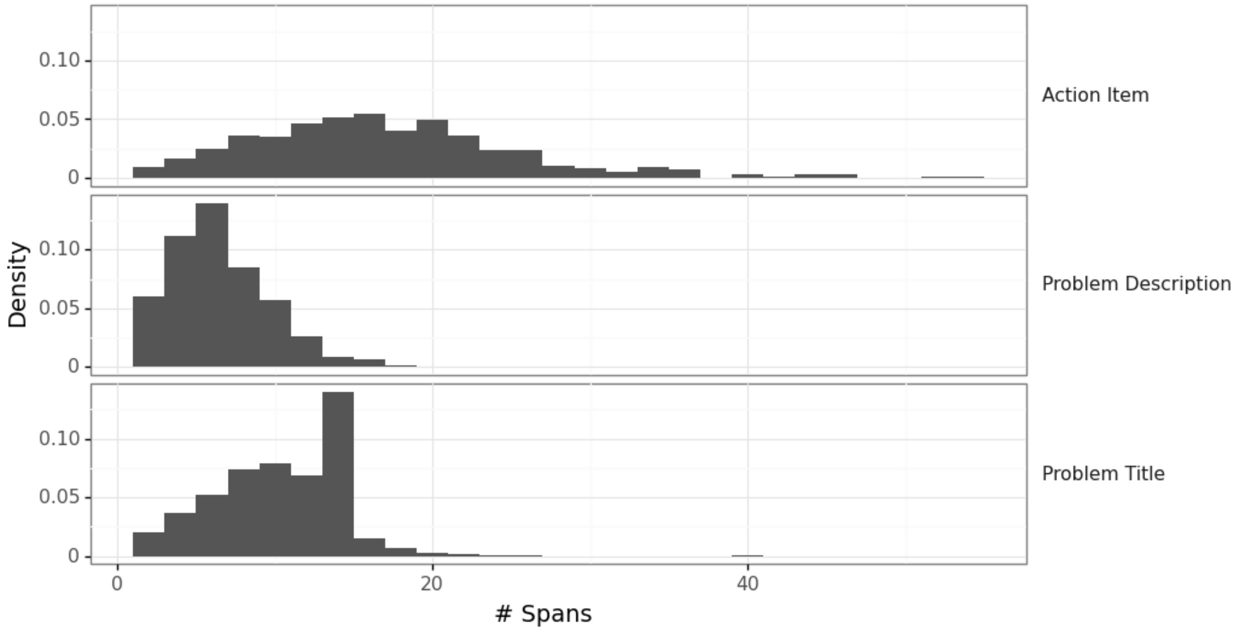


Histogram of the number of spans per note by the span type (row). The histogram is normalized by the number of spans per type. “# Spans” is the number of spans.

#### Supplementary Figure 3 - Span Type per Note Distribution by Service

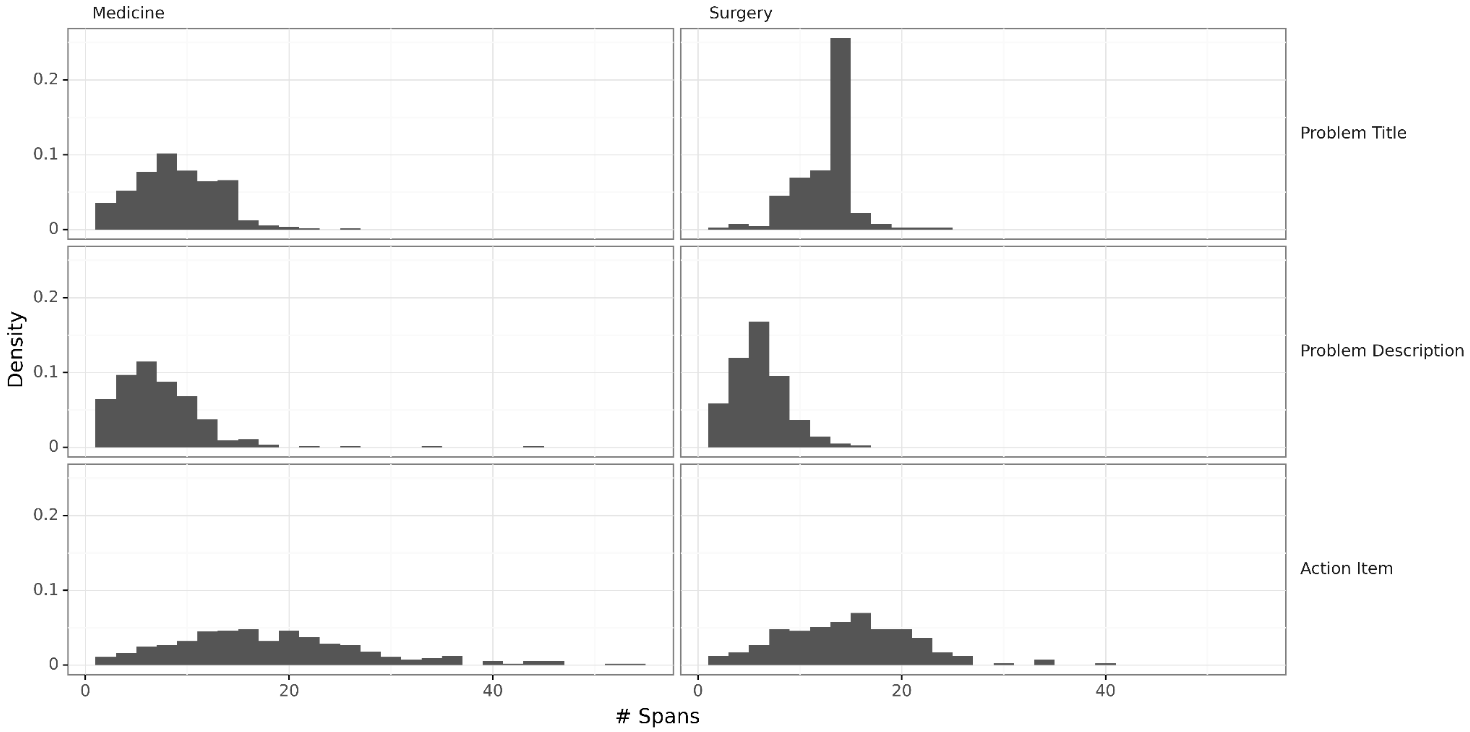


Histogram of the number of spans per note by the span type (row) and the care team service (columns).

#### Supplementary Figure 4 - Action Item Type Rater Confusion Matrix

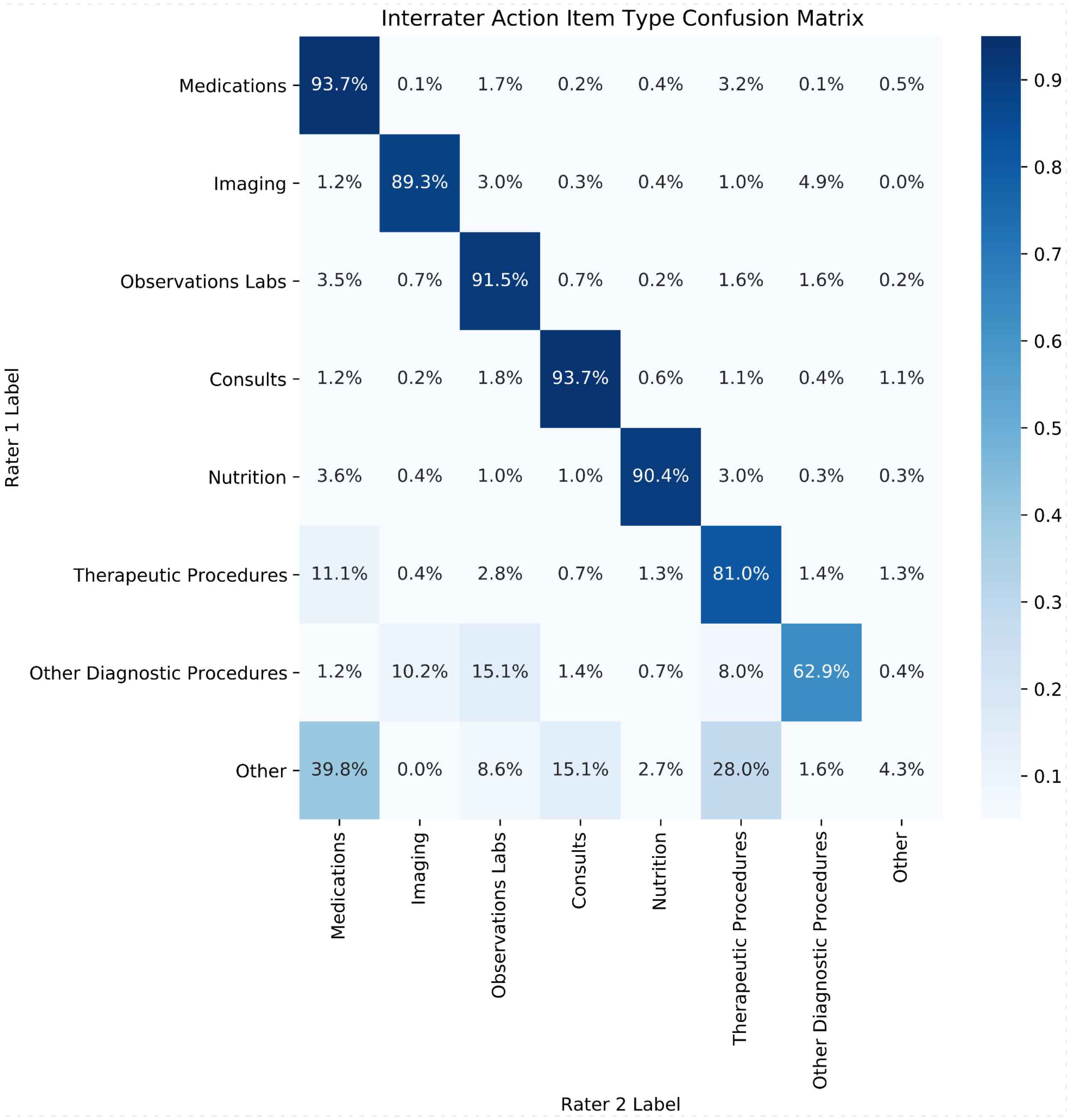


Interrater agreement for action item types as a confusion matrix of pairwise agreement. Percentage in each cell is the percent of spans labeled as the type specified by rater 1 (row) that were labeled as the type specified by rater 2 (column). For example, 1.7% of spans rated by rater 1 as Medications (first row) were labeled by rater 2 as Observations / Labs (third column). Raters are arbitrarily numbered. Cells are colored by the percentage as denoted by the colorbar to the right.

Supplementary Figure 5 - Augmentation Examples

### Base Note (No Aug.)

Assessment and Plan:

80 y/o f with a history of CHF, DM2, AFib.

Admitted with a high fever and HoTN 90/60.

Sepsis: dd pneumonia, UTI. bcx at EM neg.

- CXR am
- f/u bc, ucx
- trend wbc, lac
- Taz/Vanc.

Rhythm: hx of PAF. on metoprolol at home.

- monitor on tele
- rate control with bb

### Partially flatten

Assessment and Plan:

80 y/o f with a history of CHF, DM2, AFib.

Admitted with a high fever and HoTN 90/60

Sepsis.

  - trend wbc, lac.
  - f/u bc, ucx.
  * dd pneumonia, UTI.
  - Taz/Vanc. bcx at EM neg.
- CXR am.

### Rhythm: rate control with bb. hx of PAF. on metoprolol at home.

- monitor on tele.

### Mixed title delimiters

Assessment and Plan:

80 y/o f with a history of CHF, DM2, AFib.

Admitted with a high fever and HoTN 90/60

Sepsis:

  - Taz/Vanc.
  - trend wbc, lac.
- f/u bc, ucx.
  - dd pneumonia, UTI.
  * CXR am.
  - bcx at EM neg.

### Rhythm.

  * monitor on tele.
  * rate control with bb.
  * hx of PAF.
- on metoprolol at home.

### Mixed titles delimiters (with numbering)

Assessment and Plan:

80 y/o f with a history of CHF, DM2, AFib.

Admitted with a high fever and HoTN 90/60

Sepsis:

  - trend wbc, lac.
  - bcx at EM neg.
  - Taz/Vanc.
  - dd pneumonia, UTI.
  * CXR am.
- f/u bc, ucx.

2) Rhythm.

  * monitor on tele.
  * rate control with bb.
- on metoprolol at home.
  * hx of PAF.

### Flatten

Assessment and Plan:

80 y/o f with a history of CHF, DM2, AFib.

Admitted with a high fever and HoTN 90/60

Sepsis - f/u bc, ucx and CXR am, trend wbc, lac, bcx at EM neg
Taz/Vanc, dd pneumonia, UTI

Rhythm - rate control with bb. monitor on tele, on metoprolol at home and hx of PAF and

Data augmentations performed on assessment and plan sections. Each panel presents a randomly sampled output of the augmentation denoted by the panel title. Highlighted spans denote the active problems (purple), problem descriptions (blue) and action items (green). Action item types not shown.

#### Supplementary Figure 6 - Loss patterns

| | Ground Truth | Model |
|---|---|---|
| Missed Spans | Access: peripherals, right tunneled HD line, right port-a-cath<br>Code: full<br>Communication: Patient<br>Disposition: ICU care as patient is on pressors | Access: peripherals, right tunneled HD line, right port-a-cath<br>Code: full<br>Communication: Patient<br>Disposition: ICU care as patient is on pressors |
| | Imaging: CXR today, unchagned from prior<br>Fluids: D5 1/2 NS, heplock fluids once TF at goal<br>Consults: thoracic surgery/interventional pulmonology<br>Billing Diagnosis: | Imaging: CXR today, unchagned from prior<br>Fluids: D5 1/2 NS, heplock fluids once TF at goal<br>Consults: thoracic surgery/interventional pulmonology<br>Billing Diagnosis: |
| Confusing descriptions and action items | Cardiovascular: Aspirin, Stable hemodynamically. Colchecine and indomethacine.<br>Pulmonary: IS, Stable on RA. OOB --> [**Last Name (un) **] | Cardiovascular: Aspirin, Stable hemodynamically. Colchecine and indomethacine.<br>Pulmonary: IS, Stable on RA. OOB --> [**Last Name (un) **] |
| | Hematology: Serial Hct, Stable anemia. Monitor. No indication for PRBC transfusion<br>Endocrine: RISS, BG well controlled. Keep < 150 | Hematology: Serial Hct, Stable anemia. Monitor. No indication for PRBC transfusion<br>Endocrine: RISS, BG well controlled. Keep < 150 |
| | Endo: RISS, on home Prednisone, on Hydrocortisone. | Endo: RISS, on home Prednisone, on Hydrocortisone. |
| | Nutrition: Regular diet, - some po intake. Discuss PEG with ptnt and family due to poor caloric intake | Nutrition: Regular diet, - some po intake. Discuss PEG with ptnt and family due to poor caloric intake |
| | Infectious Disease: Check cultures, Aspergilus and Stenotrophomonas on BAL at OSH - treating with Voriconazole untill fungal BAL cultures return neg. likely d/c today | Infectious Disease: Check cultures, Aspergilus and Stenotrophomonas on BAL at OSH - treating with Voriconazole untill fungal BAL cultures return neg. likely d/c today |
| | Lines / Tubes / Drains: Foley, Chest tube x2 pleural. PICC eval. | Lines / Tubes / Drains: Foley, Chest tube x2 pleural. PICC eval. |
| Splitting or merging spans | Pulm: Monitor for pulmonary edema, may need Lasix and BiPAP as needed.<br>Albuterol and Ipratropium nebs prn.<br>GI: NPO x ice chips, PPI | Pulm: Monitor for pulmonary edema, may need Lasix and BiPAP as needed.<br>Albuterol and Ipratropium nebs prn.<br>GI: NPO x ice chips, PPI |
| | Pulmonary: Chest PT, OOB; Phys therapy and occupational Therapy. | Pulmonary: Chest PT, OOB; Phys therapy and occupational Therapy. |

Loss profiles of the model against ground truth. Each row displays a single loss profile of the model (right) compared with the ground truth (left). Highlighted spans denote the active problems (purple), problem descriptions (blue) and action items (green). Action item types are not shown.

### Supplementary Text - Labeling Instructions and Quiz

#### Background

Assessment and plan sections tend to follow a problem oriented structure. In this task we'd like to annotate the problems for assessment and their associated action items and categorize those action items. The task consists of labeling spans of free text.

#### Task

In this task you'll get a medical note. Your task is to label Assessment and Plan sections with the following types of text spans:

1. Problem titles - They can be a single condition (e.g. pneumonia, COPD ex.), contain multiple conditions (e.g. DKA/AG acidosis), a clinical development (e.g. IHD s/p CABG), a body system (e.g. GI) a clinical concept (e.g. FEN) or other patient care concepts (e.g. Prophylaxis, Access).
2. Problem descriptions - assessment of the problem, states facts about the patient's previous and current condition in the prism of the discussed problem. Does not imply an action should be done.
3. Action items - prescribed plan for the given problem. Includes both explicit and implicit actions. Action items are atomic statements which should be understandable on their own, for example in a bulleted list. For action items the task consists of both span markup as well as categorizing them to 8 categories.

#### General Guidance

Color guide - problem title, problem description, action item

| | *Explanation* | *Examples* |
|---|---|---|
| Text before any problem titles | To be ignored | "Assessment and Plan: 90M with retroperitoneal bleed after mechanical<br>fall while supratherapeutic on coumadin<br>Neurologic: Neuro checks Q: 4 hr, patient reports that fall was"<br><br>Everything before "Neurologic" is to be ignored. |
| Multiple A&P sections | All should be labeled | "**Assessment and Plan**<br>COAGULOPATHY, PAIN CONTROL (ACUTE PAIN, CHRONIC PAIN)<br>**Assessment and Plan**: 90M with retroperitoneal bleed after mechanical<br>fall while supratherapeutic on coumadin<br>Neurologic: Neuro checks Q: 4 hr, patient reports that fall was ..." |
| Length of labels | Labels will usually span a whole sentence. Usually separated by bullets, punctuation or a conjunction/conditional. consecutive labels of the same type should not be joined. | Neurologic: Neuro checks Q: 4 hr, patient reports that fall was mechanical and he denies LOC, patient has some bruising around his LEFT<br>eye; hydromorphone 0.25mg q3h prn<br><br>Color coded as problem title, problem description, action item<br>Problem description spans multiple lines |
| Span labeling | Labeling an exact span is difficult, try to include only characters that are related to the label but don't stress on accidentally captured spaces which can be automatically corrected | Neurologic: Neuro checks Q: 4 hr, patient reports that fall was... around his LEFT eye; hydromorphone 0.25mg q3h prn<br><br>Automatically corrects to:<br><br>Neurologic: Neuro checks Q: 4 hr, patient reports that fall was... around his LEFT eye; hydromorphone 0.25mg q3h prn |

#### Action item labeling

Action items are to be classified into one of the 8 categories:

#### Action item categories

| *Category* | *Description* | *Examples* |
|---|---|---|
| Medications | therapeutic procedure discussing the administration of a substance to the patient excluding nutritional plans. Includes fluids. Includes various actions related to the administration e.g. hold, stop, continue. | “Aspirin”, “continue statin”, “renally dose meds”, “hold anticoagulants”, “D5W 500cc”, “transfusions”<br><br>O2, fluids, electrolytes and blood products are all considered medications for this task. |
| Imaging | diagnostic procedure achieved via radiologic or nuclear medicine apparatuses. These include both imaging procedures performed by radiology (CT, MRI) as well as other departments (echocardiogram or ob/gyn ultrasound) | “serial cxrs”, “repeat ct head” “tte tomorrow” |
| Observations / Labs | diagnostic procedure measuring a quantity in the patient’s physiology | “monitor telemetry”, “trend creatinine” |
| Other Diagnostic Procedures | other procedures with a diagnostic intent which don’t fit imaging or observation / labs | eeg, ecg, egd |
| Therapeutic Procedures | other procedures with a therapeutic intent which are not drugs or nutritional plans. Both invasive and non-invasive procedures, both directly therapeutic and prophylactic. | surgery, hemodialysis, line placements, prophylaxis with pneumoboots, head of bed elevation. |
| Consults | both physician specialists and other caregivers. Both requests for consults and consult recommendations when those don’t entail other action item categories. Both inpatient stay consults and outpatient appointments. | “sw consult”, “appreciate id recs”.<br><br>**not** “hd tomorrow per nephro recs” (which would classify as therapeutic procedure) |
| Nutrition | nutrition plans including substances given for nutritional treatment which do not treat other concurrent abnormalities (e.g. electrolytes). Also includes hold, stop, | “tpn”, “tube feeds”, “npo”<br><br>but **not** “1/2NS+D5W” or “replete electrolytes” (which |

| | | |
|---|---|---|
| | continue of said plans. | would classify as meds). |
| Other | action items which don't clearly fit any other category, please explain in category comment and reach out for discussion. | |

##### Additional instructions for action items:

| | *Explanation* | *Examples* |
|---|---|---|
| Multiple actions per action item | Action items should be split if they are understandable on their own. if additional "modifiers" are shared across multiple actions, label them together. | "continue vanco, cefepime for 10 days (today = day #5)" all are related to both "continue" and "for 10 days" |
| | | "FEN: IVF, replete electrolytes, NPO after midnight" contains 3 separate action items (in yellow). In the "FEN" example, while IVF and replete electrolytes are both "medication" they don't have any modifiers to connect them and thus should be considered separate. |
| | | "f/u blood, urine cultures" is a single action item as "f/u blood" is meaningless without denoting "cultures" |
| Conditional and temporal relationships | should be treated as separate action items when each stands on its own. | "If worsening abdominal exam/sepsis, increasing lactate; would obtain abdominal CT for evaluation of toxic megacolon or surgical disease"<br><br>Should be split as "If worsening abdominal exam/sepsis, increasing lactate" (observations/labs),<br><br>"would obtain abdominal CT for evaluation of toxic megacolon or surgical disease" (imaging) |
| | | "Follow lytes and replete aggressively" - should not be split as "replete aggressively" doesn't stand on its own. |
| | | "restart home HCTZ during hospitalization if BPs can tolerate" should not be split as the condition ("if BPs can tolerate") cannot stand on its own. |
| Joint action item and problem description | When an action item contains information about the state of the patient which modifies the action item and would otherwise be classified as problem | As for the example above, "will have anesthesia present, may discuss with surgery also, given extreme difficulty of intubation in the ED" - "given extreme difficulty of intubation in the ED" is a modifiers of the action item and thus should be marked as part of the action item. |

| | | |
|---|---|---|
| | description, the text containing the description should be left as part of the action item. | "Was on heparin gtt on the floor but will d/c now given hct drop." similarly an action item though the description is before the action item. |
| Interleaving action items and problem description | problem description can come after action items. They are differentiated from one another by having no action intent, implicitly or explicitly | # Hypercarbic Respiratory Failure: likely due to CNS depression after multiple ingestions.<br>- on pressure support [**5-14**] with good sats<br>- wean sedation<br>-cuff leak present this morning<br>-increased secretions this morning<br>-possible extubation later today or tomorrow… |
| Differentiating descriptions from action items | Some may be more subtle. An action item containing only an object can refer to both a state or an action. In cases where it is unclear from the context which to choose, prefer action items | "# Access: PICC line, PIV<br># Communication: wife, daughter (phone numbers in chart)"<br>the communication problem contains only problem description (and title) as "wife" refers to "wife is the point of contact" and not "call wife". For access it is unclear whether "PICC line" and "PIV" are orders ("insert PICC line") or the current state ("has PICC line"), thus we prefer action items in this case. |
| No action is a problem description | Denoting a lack of actions needed for a problem is a problem description. | Hematology:<br>--No labs<br>Imaging: none<br>Fluids: None<br>Lines / Tubes / Drains: PIV, ETT, Foley, A-line is currently not required |

#### Quiz

Answers are in bold

| Question | Text | Options: |
|---|---|---|
| 1. **Problem Titles**: in the following text, select all “problem titles”: | Assessment and Plan:<br>50 y/o m with a hx of Asthma.<br>Admitted for SOB.<br># Asthma. ...<br># ID: ...<br># FEN: ...<br># Prophylaxis: ...<br># Access: ...<br>Contact: ...<br>Code: ...<br>Dispo: ... | 1. 50 y/o m with a hx of Asthma.<br>2. Admitted for SOB.<br>**3. Asthma**<br>**4. ID**<br>**5. FEN**<br>**6. Prophylaxis**<br>**7. Access**<br>**8. Contact**<br>**9. Code**<br>**10. Dispo** |
| 2. Indicate which spans of text represent problem titles, problem descriptions, and/or action items | # ESRD- on HD MWF - continue HD, replete lytes | 1. ESRD **- PT**<br>2. on HD MWF **- PD**<br>3. continue HD **- AI**<br>4. replete lytes **- AI** |
| 3. Indicate which spans of text represent problem titles, problem descriptions, and/or action items | RENAL: making good urine, creatinine 0.5, will watch for DI, follow<br>sodium. Will try to keep even fluid control. | 1. RENAL **- PT**<br>2. making good urine **- PD**<br>3. creatinine 0.5 **- PD**<br>4. will watch for DI **- AI**<br>5. follow sodium **- AI**<br>6. Will try to keep even fluid control. **- AI** |
| 4. **Problem Description /Action Item splitting** - what are the correct set of problem descriptions and action items? | Pulmonary: Once family makes decision, extubate, morphine/ativan prn. | 1. “Once family makes decision, extubate, morphine/ativan prn.” (1 action item total)<br>2. “Once family makes decision” (PD), “extubate” (AI), “morphine/ativan prn.” (AI) (3 total)<br>**3. “Once family makes decision, extubate” (AI), “morphine/ativan prn.” (AI) (2 total)** |
| 5. **Problem Description /Action Item splitting** - what are the correct set of problem descriptions and action items? | Acute respiratory failure: Now extubated. Diuresed overnight, will give morphine prn. Cont vanc/cefepime. Will continue to keep him in the ICU. | 1. “Now extubated. Diuresed overnight” (PD), “will give morphine prn” (AI), “Cont vanc/cefepime” (AI), “Will continue to keep him in the ICU” (PD) (4 total)<br>2. “Now extubated. Diuresed overnight” (PD), “will give |

| | | |
|---|---|---|
| | | morphine prn" (AI), "Cont vanc/cefepime" (AI), "Will continue to keep him in the ICU" (AI) (4 total)<br>3. "Now extubated" (PD), "Diuresed overnight, will give morphine prn" (AI), "Cont vanc/cefepime" (AI), "Will continue to keep him in the ICU" (AI) (4 total)<br>4. "Now extubated. Diuresed overnight" (PD), "will give morphine prn" (AI), "Cont vanc" (AI), "cefepime" (AI), "Will continue to keep him in the ICU" (PD) (5 total)<br>**5. "Now extubated. Diuresed overnight" (PD), "will give morphine prn" (AI), "Cont vanc" (AI), "cefepime" (AI), "Will continue to keep him in the ICU" (AI) (5 total)** |
| 6. **Action Item splitting** - what are the correct set of action items? | Prophylaxis: PPI, heparin sq, bowel regimen. | **1. "PPI", "heparin sq", "bowel regimen" (3 total)**<br>2. "PPI, heparin sq, bowel regimen" (1 total). |
| 7. **Action Item splitting** - what are the correct set of action items? | f/u sputum, blood, urine cultures | **1. "f/u sputum, blood, urine cultures" (1 total)**<br>2. "f/u sputum", "blood", "urine cultures" (3 total) |
| 8. **Action Item splitting** - what are the correct set of action items? | cont. levophed for BP support & wean as tolerated | **1. "cont. levophed for BP support & wean as tolerated" (1 total)**<br>2. "cont. levophed for BP support", "wean as tolerated" (2 total) |
| 9. **Action Item type** - select the correct action item type for the marked action item | FEN: **IVF D5W 50cc/hr** | **Medication**, Imaging, Observations/Labs, Consults, Nutrition, Therapeutic Procedures, Other Diagnostic Procedures, Other |
| 10. **Action Item type** - select the correct action item type for the marked action item | ID: Follow cultures, continue antibiotics, goal CVP 8-12, **pressors for BP support as needed, goal MAP >60**. | **Medication**, Imaging, Observations/Labs, Consults, Nutrition, Therapeutic Procedures, Other Diagnostic Procedures, Other |
| 11. **Action Item type** - select the correct action item type for the marked action item | CVS: known AS, **TTE today** | Medication, **Imaging**, Observations/Labs, Consults, Nutrition, Therapeutic Procedures, Other Diagnostic Procedures, Other |

| | | |
|---|---|---|
| 12. **Action Item type** - select the correct action item type for the marked action item | Resp distress: Intubated, **wean O2 as possible** | **Medication**, Imaging, Observations/Labs, Consults, Nutrition, Therapeutic Procedures, Other Diagnostic Procedures, Other |
| 13. **Action Item type** - select the correct action item type for the marked action item | Anemia: monitor Hct daily, **transfuse for Hct<21%** | **Medication**, Imaging, Observations/Labs, Consults, Nutrition, Therapeutic Procedures, Other Diagnostic Procedures, Other |
